# Identifying patients requiring treatment for depression in the postpartum period from common electronic medical record data available antepartum using machine learning

**DOI:** 10.1101/2022.11.30.22282941

**Authors:** Colin Wakefield, Martin G. Frasch

## Abstract

**Objective:** Depression requiring treatment in the postpartum period (PPD) significantly impacts maternal and neonatal health. While preventive management of depression in pregnancy has been shown to decrease the negative effects, current methods for identifying at-risk patients are insufficient. Given the complexity of the diagnosis and interplay of clinical/demographic factors, we tested if machine learning (ML) techniques can accurately identify patients at risk of PPD.

**Study Design:** This is a retrospective cohort study of the NIH Nulliparous Pregnancy Outcomes Study (nuMoM2b) which enrolled 10,038 nulliparous people. The primary outcome was PPD. We constructed and optimized four ML models using distributed random forest modeling based on the nuMoM2b dataset. Model 1 utilized only readily obtainable sociodemographic data. Model 2 added maternal pre-pregnancy mental health data. Model 3 utilized recursive feature elimination to construct a parsimonious model. Model 4 further titrated the input data to exclude pre-pregnancy mental health variables.

**Results:** Of 8454 births, 338 (4%) were complicated by PPD. Model 3 was the highest performing, demonstrating the area under the receiver operating characteristics curve (AUC) of 0.91(±0.02). Models 1-3 identified the 9 variables most predictive of depression hierarchically ranging from BMI (highest), prior depression, age, income, medications, education, past medical history, race, and prior anxiety (lowest). In model 4, the AUC remained at 0.80(±0.04).

**Conclusions:** Counterintuitively, the presence of pre-pregnancy mental health conditions is not the most predictive factor of PPD. Furthermore, PPD can be predicted with high accuracy for individual patients using antepartum information commonly found in the EMR.

## Introduction

In the United States, as many as 1 in 7 pregnant people will develop perinatal depression in the postpartum period (PPD)^1^. Yet, only 50% of affected women are diagnosed and even less receive appropriate treatment^2^. Often underdiagnosed and undertreated due to stigma and mislabeling as the “mommy blues”, failing to recognize the impact of untreated PPD significantly affects both mother and child.

PPD impacts patients differently depending on the time of onset. Prepartum onset increases risk of preterm birth, preeclampsia, low birth weight, maternal suicide, and poor cognitive and behavioral trajectories in the fetus^3^. The maternal suicide risk associated with prepartum onset should not be underestimated as it surpasses both hemorrhage and hypertensive disorders as the leading cause of maternal mortality during pregnancy^4^. PPD carries the additional risk of negatively impacting child safety due to depressive symptoms in the mother. Postpartum presentation has been linked to less breastfeeding, poor adherence to infant safety recommendations, fewer well-child visits, and an overall increased risk of child neglect. Mothers during this period are less likely to care for their own needs and have a pronounced risk of self-harm as suicide accounts for 20% of deaths in moms within the year following birth^1^.

The long-term consequences of PPD are often underestimated. Feelings of intense sadness, anxiety, and anhedonia impede bonding with the infant, resulting in cognitive, behavioral, and emotional developmental delays for the child as well as delays in social and communication skills^1^. Such delays can have lifelong ramifications for the child and pose a real threat towards their capacity for social adjustment.

Fortunately, early detection and subsequent preventive interventions are highly efficacious in mitigating the consequences of PPD. In 2019, the US preventive service task force (USPSTF) conducted a meta-analysis, finding that interpersonal therapy and/or cognitive behavioral therapy (CBT) in at-risk pregnant people is associated with a 39% reduction in the likelihood of developing PPD. In specific at-risk populations, the associated risk reductions are as high as 53%^5^.

Even though the impacts of PPD can be profound and early intervention has been shown to significantly reduce incidence, there are currently no tools available for easy identification of at-risk patients. The lack of effective screening tools is a public health issue as early interventions in at-risk people can minimize the morbidity and mortality of PPD. The development of a tool to stratify PPD risk and assist with resource allocation to in-need populations has the potential to greatly improve the perinatal health care delivery.

As such, using machine learning models (ML), attempts have been made with minimal success to develop predictive models to identify patients at risk of PPD. Over the past ten years, there have been 13 publications presenting ML models to predict PPD. A common issue among these models has been the failure to accuratley predict PPD using only readily obtainable data. Previous models have attempted to incorporate input data that is not uniform in nature nor easily collected, such as psychological resilience, dental hygiene, and 5-HTT-GC levels ^6–8^. Prior models have routinely incorporated late gestation or postpartum data, greatly reducing their predictive value. A substantial number of PPD cases arise early in pregnancy (1st trimester: 7%, 2nd trimester: 13%, 3rd trimester: 12%), thus models relying on late gestation/postpartum data fail to capture substantial patient populations ^9^. Moreover, in order for PPD prediction models to be effective, at-risk patients must be identified early enough in pregnancy for cognitive-behavioral therapy to take effect. Therefore, a PPD prediction model needs to use input data that can be easily gathered early in or prior to pregnancy; something only one prior model has attempted with modest success (68% model sensitivity)^10^.

Given the morbidity and mortality of PPD, there is a clear need for effective screening tools to quantify individual patient risk for PPD. The complexity of the diagnosis and interplay of clinical/demographic factors make it difficult for clinicians to individually identify at-risk patients. Therefore our team tested if ML techniques can accurately identify patients at risk of PPD using only pre-pregnancy data found in the electronic medical records.

## Methods

To test it ML techniques can accurately identify patients at risk of PPD requiring treatment, our team performed a retrospective cohort analysis of the “Nulliparous Pregnancy Outcomes Study: monitoring mothers-to-be (nuMoM2b)” dataset. The NuMoM2b study is a prospective cohort study administered by the NIH in which 10,038 nulliparous women with singleton pregnancies were enrolled from geographically-diverse hospitals affiliated with 8 clinical centers. It is unique in that it was designed to investigate the interrelated mechanisms of common adverse pregnancy-related outcomes. Through its design, the nuMoM2b study subsequently created one of the largest public datasets from which the relationship between hundreds of sociodemographic variables and different pregnancy-related outcomes can be investigated. The data can be obtained via the NICHD DASH portal.^11^

Full details of the study protocol have been published previously.^12,13^ In brief, women were eligible for enrollment if they had a viable singleton gestation, had no previous pregnancy that lasted more than 20 weeks of gestation, and were between 6 weeks 0 days’ gestation and 13 weeks 6 days’ gestation at recruitment. Exclusion criteria were: maternal age less than 13 years, history of 3 or more spontaneous abortions, current pregnancy complicated by a suspected fatal fetal malformation, known fetal aneuploidy, assisted reproduction with a donor oocyte, multifetal reduction, or plan to terminate the pregnancy. Patients also were excluded if they were already participating in an intervention study anticipated to influence pertinent maternal or fetal outcomes, were previously enrolled in the nuMoM2b study, or were unable to provide informed consent. A common protocol and manual of operations were used for all aspects of the study. Each site’s local governing Institutional Review Board approved the study and all women provided informed written consent prior to participation.

Participant data were collected by trained research personnel during three antepartum study visits. These visits were scheduled to occur between 6 weeks 0 days’ and 13 weeks 6 days’ gestation (Visit 1), 16 weeks 0 days’ and 21 weeks 6 days’ gestation (Visit 2), and 22 weeks 0 days’ and 29 weeks 6 days’ gestation (Visit 3). A woman’s self-identified race and ethnicity was categorized as non-Hispanic white, non-Hispanic black, Hispanic, Asian or “other”. At least 30 days after delivery, trained and certified chart abstractors reviewed the medical records of all participants and recorded final maternal and birth outcomes.

The list of predictor variables used in the present ML models is provided in **Table 1**.

**Table 1.**
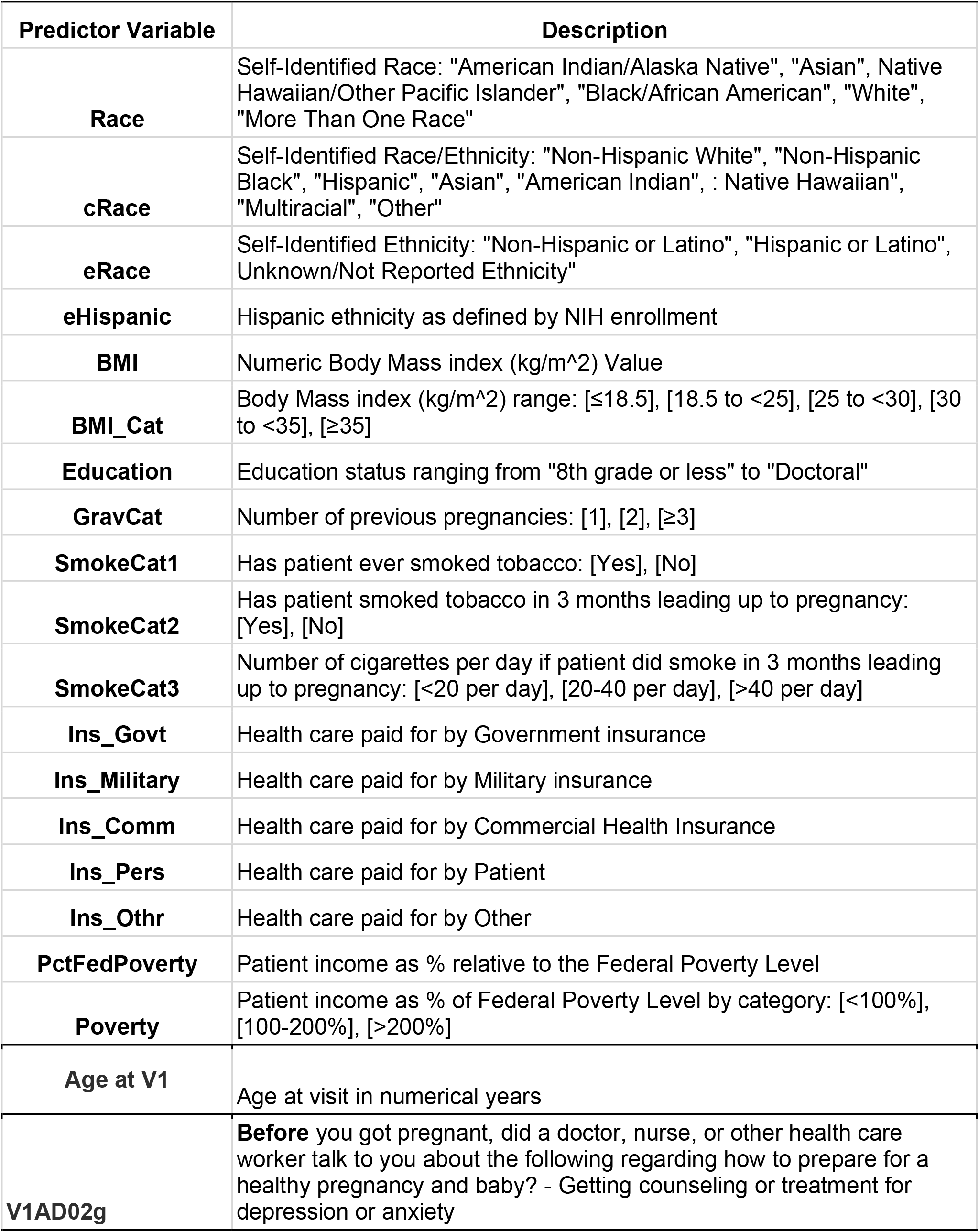
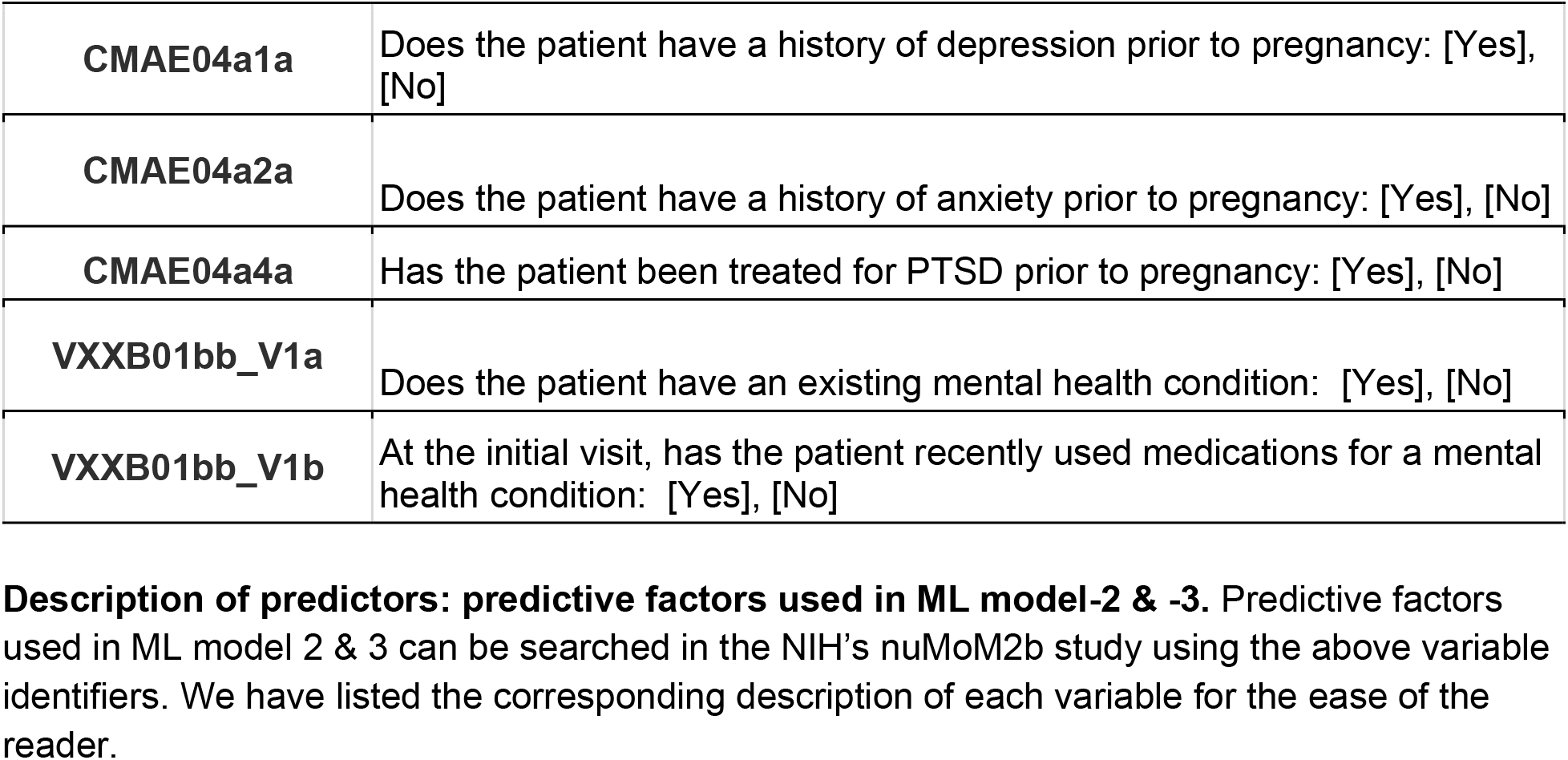
Description of predictor variables.

| **Predictor Variable** | **Description** |
| --- | --- |
| **Race** | Self-Identified Race: "American Indian/Alaska Native", "Asian", Native Hawaiian/Other Pacific Islander", "Black/African American", "White", "More Than One Race" |
| **cRace** | Self-Identified Race/Ethnicity: "Non-Hispanic White", "Non-Hispanic Black", "Hispanic", "Asian", "American Indian", : Native Hawaiian", "Multiracial", "Other" |
| **eRace** | Self-Identified Ethnicity: "Non-Hispanic or Latino", "Hispanic or Latino", Unknown/Not Reported Ethnicity" |
| **eHispanic** | Hispanic ethnicity as defined by NIH enrollment |
| **BMI** | Numeric Body Mass index (kg/m<sup>2</sup>) Value |
| **BMI_Cat** | Body Mass index (kg/m<sup>2</sup>) range: [≤18.5], [18.5 to <25], [25 to <30], [30 to <35], [≥35] |
| **Education** | Education status ranging from "8th grade or less" to "Doctoral" |
| **GravCat** | Number of previous pregnancies: [1], [2], [≥3] |
| **SmokeCat1** | Has patient ever smoked tobacco: [Yes], [No] |
| **SmokeCat2** | Has patient smoked tobacco in 3 months leading up to pregnancy: [Yes], [No] |
| **SmokeCat3** | Number of cigarettes per day if patient did smoke in 3 months leading up to pregnancy: [<20 per day], [20-40 per day], [>40 per day] |
| **Ins_Govt** | Health care paid for by Government insurance |
| **Ins_Military** | Health care paid for by Military insurance |
| **Ins_Comm** | Health care paid for by Commercial Health Insurance |
| **Ins_Pers** | Health care paid for by Patient |
| **Ins_Othr** | Health care paid for by Other |
| **PctFedPoverty** | Patient income as % relative to the Federal Poverty Level |
| **Poverty** | Patient income as % of Federal Poverty Level by category: [<100%], [100-200%], [>200%] |
| **Age at V1** | Age at visit in numerical years |
| **V1AD02g** | **Before** you got pregnant, did a doctor, nurse, or other health care worker talk to you about the following regarding how to prepare for a healthy pregnancy and baby? - Getting counseling or treatment for depression or anxiety |
| **CMAE04a1a** | Does the patient have a history of depression prior to pregnancy: [Yes], [No] |
| **CMAE04a2a** | Does the patient have a history of anxiety prior to pregnancy: [Yes], [No] |
| **CMAE04a4a** | Has the patient been treated for PTSD prior to pregnancy: [Yes], [No] |
| **VXXB01bb_V1a** | Does the patient have an existing mental health condition: [Yes], [No] |
| **VXXB01bb_V1b** | At the initial visit, has the patient recently used medications for a mental health condition: [Yes], [No] |

**Description of predictors: predictive factors used in ML model-2 & -3.** Predictive factors used in ML model 2 & 3 can be searched in the NIH's nuMoM2b study using the above variable identifiers. We have listed the corresponding description of each variable for the ease of the reader.

The analyses have been performed in R 4.1.1 with h2o 3.34.0.3 (computationally efficient ML modeling framework). The training was done on 80% of the data (10-fold); validation was performed on the 20% of the held-out data and the model performance is reported from that unseen validation dataset. The code, results, and the <u>notebook</u> for validation and extension of the presented solution have been deposited as open-source.^14^ The following results are reported for the Distributed random forest ML model and were similar for Decision Tree, Naïves Bayes Classifier and AutoML approaches, all reported in the shared notebook.

## Results

An ML titration approach was deployed to 1) optimize performance and 2) maximize model practicality by minimizing the number of variables included. Of the 10,038 women enrolled in this prospective cohort, 9,470 were eligible for the present analysis following exclusion for lack of pregnancy outcome data, lack of race-ethnicity data, and pregnancy terminations/fetal death. Among eligible women, 5,721 (60.4%) were non-Hispanic white, 1,307 (13.8%) were non-Hispanic black, 1,586 (16.7%) were Hispanic, 379 (4.0%) were Asian, and 477 (5.0%) were of another race or ethnicity. Following further exclusion due to a lack of follow-up visits, 8545 participants’ data was available for the ML training to predict PPD which was reported in 338 cases.

Titration began with model 1, taking a simple approach that only included readily obtainable sociodemographic data (insurance, income, education, race, BMI, smoking status) and two pre-pregnancy mental health status variables (prior post-traumatic stress disorder (PTSD) & prior discussion of mental health treatment with a provider). Model 1 performed with an AUC of 0.656 (± 0.09) and predictive accuracy of 0.86 (± 0.13) (**Table 2**). Interestingly, adding pre-partum hypertension and diabetes mellitus information did not improve model performance. In model 2, another extreme was tested: use all relevant pre-pregnancy mental health variables available in the nuMoM2b dataset. The inclusion of 111 variables in model 2 considerably improved the predictive accuracy: 0.96 (± 0.01) and AUC 0.93 (± 0.02)(**Table 2**). Model 3 was derived by pruning model 2 to only include the nine most contributing variables: BMI, history of depression prior to pregnancy, age, income, recent medication use for mental health condition, education, having an existing mental health condition, race, and history of anxiety prior to pregnancy.

**Table 2:** ML model overview.

| Model | Variables | AUC (± SD) | Accuracy (± SD) |
| --- | --- | --- | --- |
| Model 1 | 6 Sociodemographic variables (*insurance, income, education, race, BMI, smoking status*) and 2 pre-pregnancy mental health variables (*prior PTSD & prior discussion on mental health treatment*) | 0.66 (± 0.09) | 0.86 (± 0.13) |
| Model 2 | All relevant pre-pregnancy data in nuMoM2b study (111) | 0.93 (± 0.02) | 0.96 (± 0.01) |
| Model 3 | 9 highest contributing variables | 0.91 (± 0.02) | 0.95 (± 0.02) |
| Model 4 | Removing mental health prior to pregnancy variables from model 3 | 0.80 (± 0.04) | 0.93 (± 0.02) |

**Predicting PPD requiring treatment: overview of model 1-4.** Distributed random forest machine learning (ML) model to predict PPD. The variables used in model 1-4 were identified using the titration approach outlined in the methods section.

Pruning to yield model 3 resulted in an overall best-performing model with a predictive accuracy of 0.95 (± 0.02) and AUC 0.91(± 0.02)(**Table 2**). Model 4 further titrated model 3 to exclude pre-pregnancy anxiety and depression, leaving the seven most contributing variables identified in model 3: BMI, age, income, recent medication use for a mental health condition, education, having an existing mental health condition, and race. Model 4 yielded a predictive accuracy of 0.93 (± 0.02) and AUC 0.80 (±0.04)(**Table 2**).

## Discussion

PPD significantly impacts maternal and neonatal health as well as the family as a whole. Affected mothers are at an increased risk of child neglect, marital conflict, mood disorder development, and poor attachment to the infant. Children born to mothers with PPD demonstrate delays in cognitive, psychosocial, motor skills, and social skills development. More importantly, PPD is a significant risk factor for infanticide and suicide, with suicide being the leading cause of maternal death in the initial postpartum year^15^. No individualized approaches currently exist to identify mothers at risk for PPD.

We present ML models that identify patients at high risk for developing PPD requiring treatment, using readily available pre-pregnancy information from the EMR.

Of the four ML models constructed, model 3 was the best performing, correctly identifying PPD requiring treatment in 91% of patients using 9 factors related to medical history & sociodemographics (**Table 2 & Figure 1**). Moreover, all 9 factors (*BMI, depression prior to pregnancy, age, income, recent medication use, education, having an existing mental health condition, race, anxiety prior to pregnancy*) are easily obtained prior to pregnancy, an important consideration when developing a clinical tool intended to provide care providers proper time for detection and intervention (**Figure 2**). Previous groups have developed predictive models for PPD but failed to use exclusively pre-pregnancy or even early pregnancy data, thus limiting the therapeutic potential of such approaches. This is because preventive intervention for PPD is largely through CBT, an evidence-based longitudinally time-intensive exercise intended to modify patients’ approach to processing stress^16^. Successful implementation of CBT can decrease PPD incidence by up to 53%^5^. However, patients identified as being at risk for developing PPD requiring treatment need adequate time to participate in CBT and develop healthy coping strategies prior to disease onset. Therefore, to maximize the therapeutic potential of a predictive model for PPD, prepregnancy data should be used.

**Figure 1.**
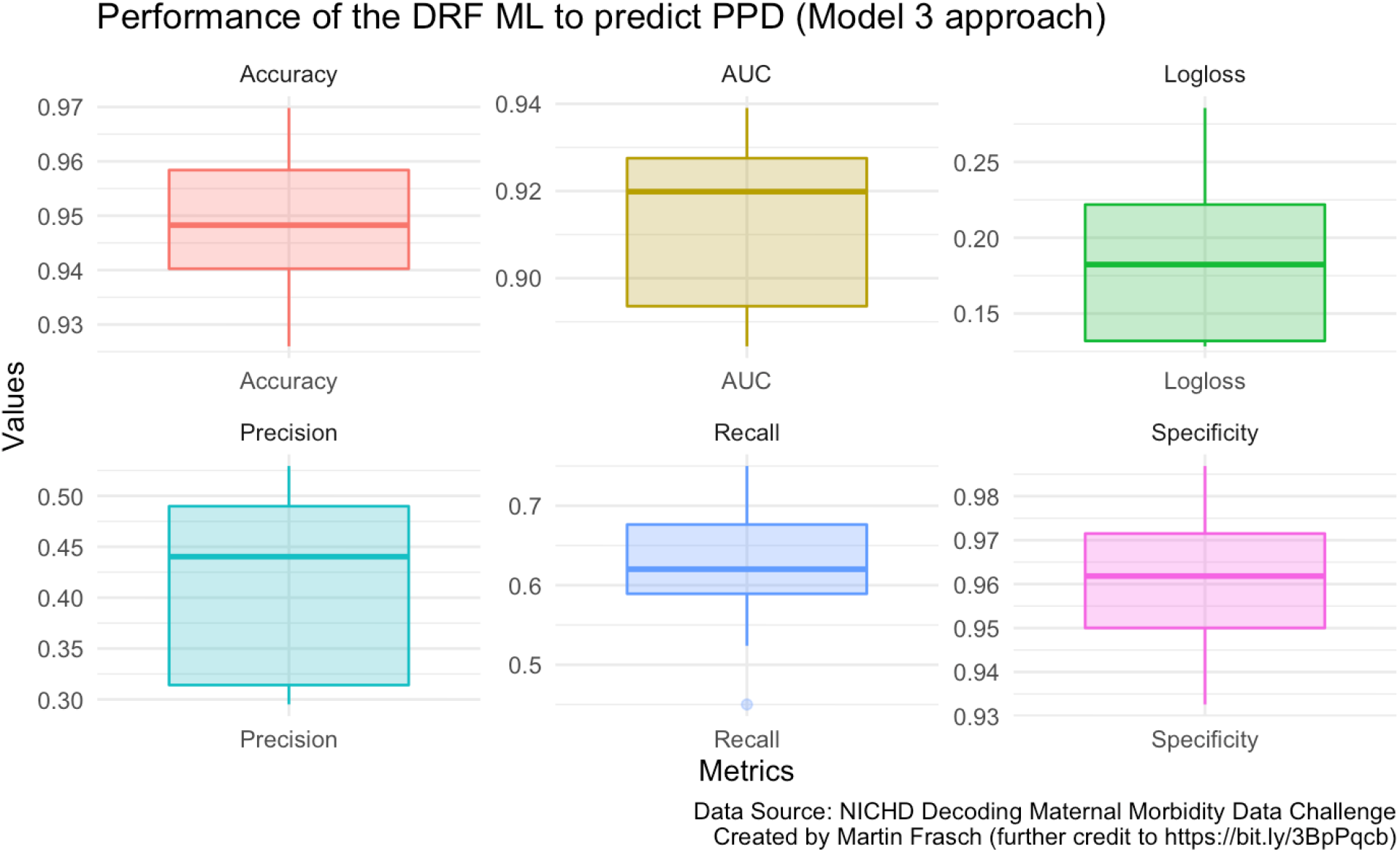
ML model performance. **ML Model 3**. Model 3 accurately identified 95% of PPD cases. AUC compares the relationship between true positives to false positives for a predictive model. AUC scores of 1.0 are considered perfect, model 3 achieved an AUC of 0.91 which is considered very strong. Logloss reflects the divergence between the predictive probability of PPD and the true rate of PPD. A logloss of 0 indicates perfect predictions, model 3 achieved a logloss of 0.18 which is considered strong. Model 3 achieved a precision of 0.44, reflecting the rate of true positives. Model 3 achieved a recall of 0.62, reflecting the rate of PPD cases correctly identified. Model 3 achieved a specificity of 0.96.

**Figure 2.**
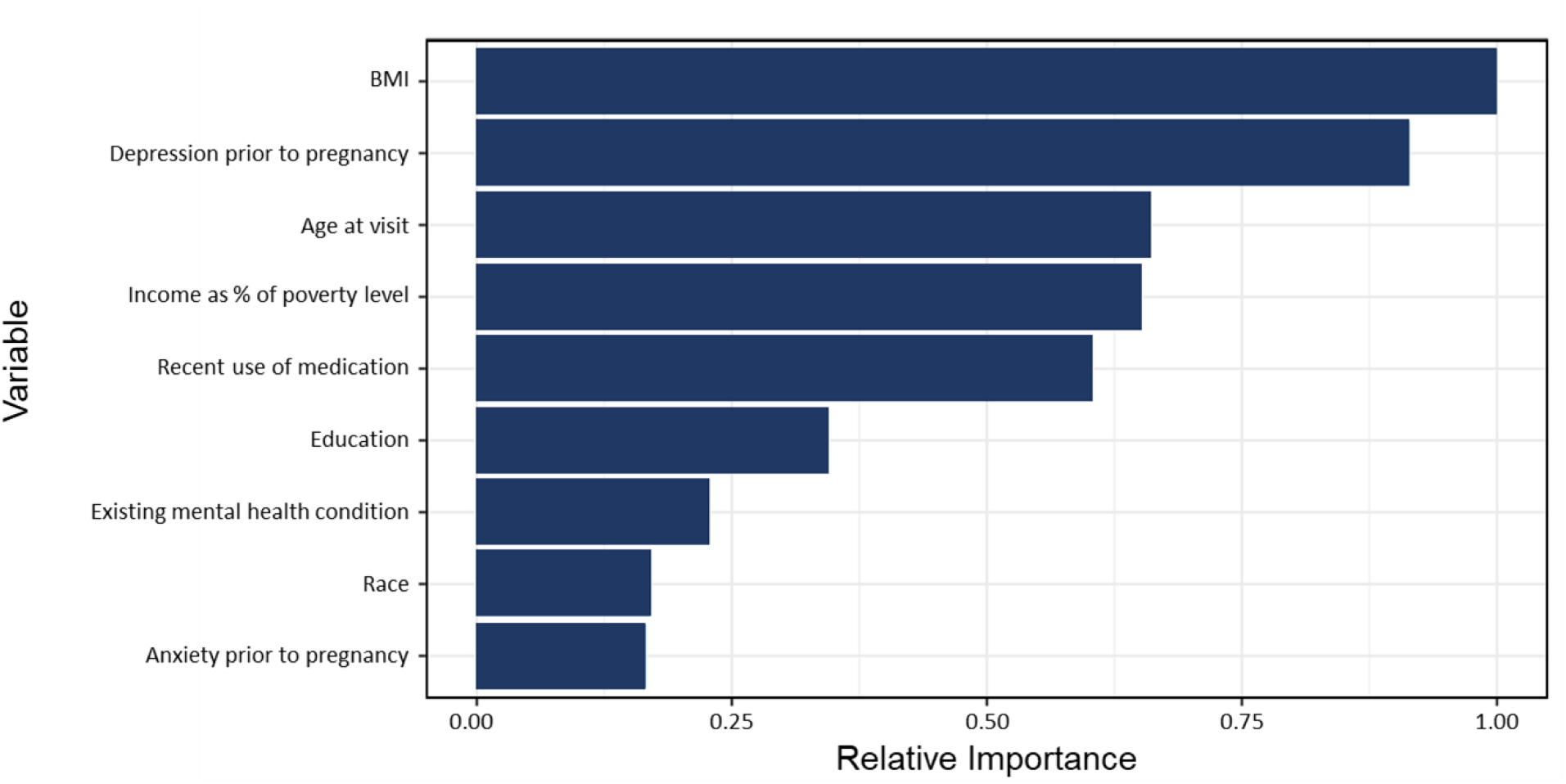
Feature importance. **Contribution of the individual factors in the prediction**. The 9 most contributing factors to the development of PPD requiring treatment, listed in descending order of relative importance: BMI, having depression during pregnancy, age at visit, patient income as a percent of the federal poverty level, recent use of medication, education status, presence of existing mental health condition, race, having anxiety prior to pregnancy.

Our models address this need using exclusively pre-pregnancy data. The sociodemographic and mental health history data points used by our ML algorithms are binary/categorical in nature and routinely found in EMRs. This approach allows for simpler integration into the modern healthcare model by limiting the need for additional data collection during patient visits. The ML code can be integrated into EMR systems, automatically pulling information gathered from intake forms and prior visits. Thus, clinicians can be provided with individual patient risk stratifications by manipulating data currently contained in the EMR.

A goal of this work was to further delineate the role of various factors in the development of PDD requiring treatment. To achieve this, model 4 was constructed by excluding maternal prepregnancy depression and anxiety history from model 3. The approach provided clarity on the role of prepregnancy mental health history in the development of PPD requiring treatment. Interestingly, model 4 performed relatively well, predicting 80% of cases while only using 7 predictors (*BMI, age, income, recent medication use, education, having an mental existing health condition, and race*) (**Table 2**). Overall, all four models identified BMI as the most contributing factor in the development of PPD requiring treatment, likely reflecting the physical, cognitive, and social impacts of body weight on human health. Depression before pregnancy is the second largest contributor to the development of PPD requiring treatment, substantiating the well-known role of prior depression in PPD. However, the sensitivity observed in model 3 was maintained in model 4 when prepregnancy psychiatric histories of depression and anxiety were excluded. **Figure 3** (ROC) demonstrates how predictive accuracy is unchanged from model 3 to model 4. This data counterintuitively highlights the relative importance of sociodemographic factors such as BMI, age, and income as greater than past psychiatric histories of anxiety & depression in the development of PPD requiring treatment.

**Figure 3.**
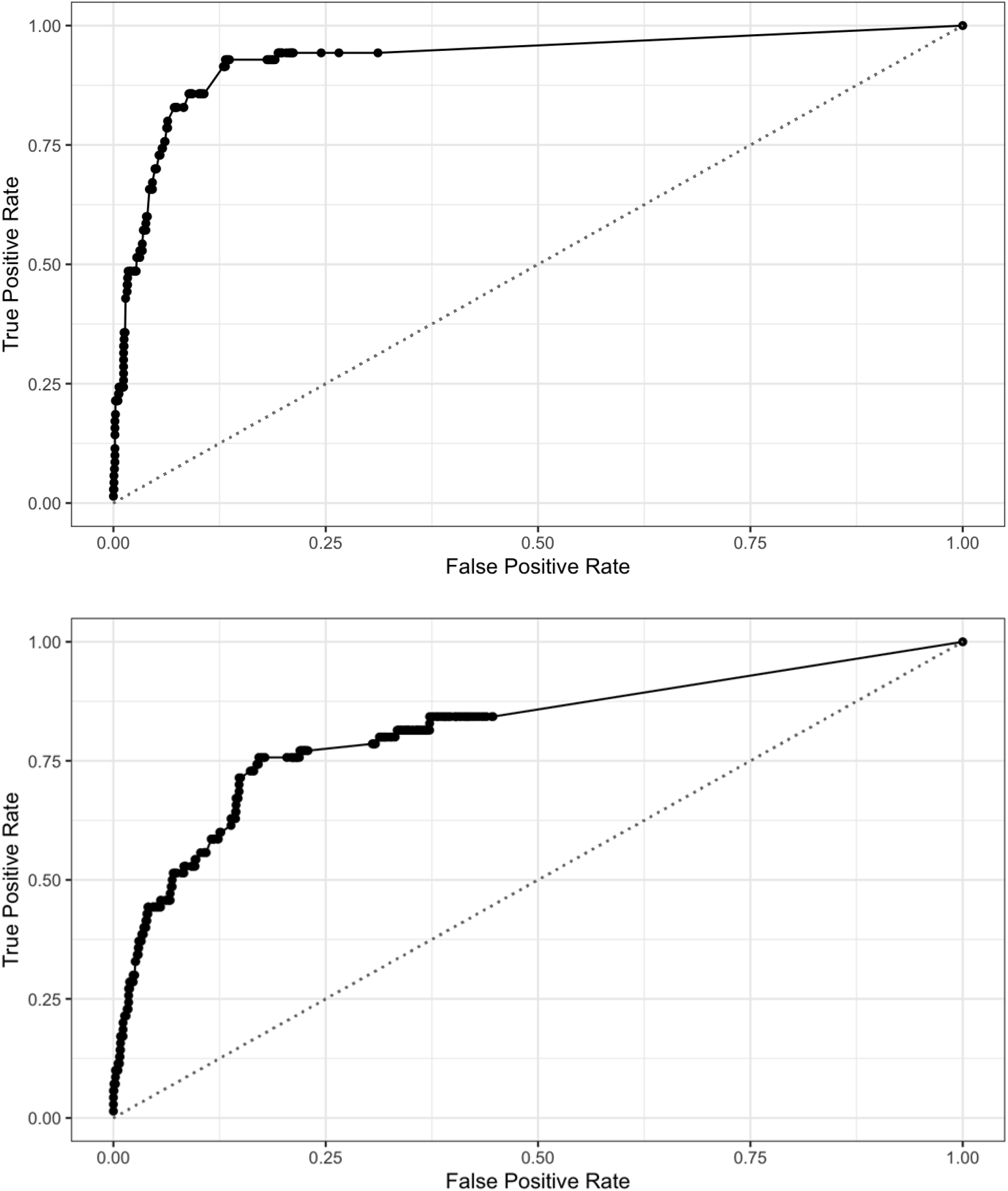
ROC performances in models 3 and 4. **Performance sustained when pruning from model 3 to model 4**. Model 3 was constructed using the 9 most contributing factors to PPD prediction. Model 4 was constructed from the 7 sociodemographic variables identified in model 3, with prepregnancy anxiety & depression excluded. The performance was largely maintained through this exclusion.

The discovery that sociodemographic factors play a pivotal role in the pathogenesis of PPD requiring treatment is key for the formulation of public health measures to target PPD. The present work provides clear directions for policymakers to decrease the incidence of PPD. Our findings highlight 3 out of the 6 most contributing factors to PPD development, BMI, income, and education, which are all modifiable through socioeconomic interventions. Based on these findings policies aimed at improving income and education equity as well as access to affordable and healthy diet have the potential to decrease the morbidity and mortality associated with PPD.

This study also provides a clear groundwork for clinicians to mitigate the impacts of PPD. Population-based studies have linked BMI to PPD, however, the relative importance of BMI in PPD pathogenesis compared to other contributing factors is not well documented^17^. Our data suggests that clinical interventions to optimize BMI are some, if not the most important steps to decrease patients’ risk of developing PPD requiring treatment. Lifestyle modifications as well as assisting patients in navigating social programs related to job training, education access, and Supplemental Nutrition Assistance Programs (SNAP) are all potential actions for clinicians to guide patients and help decrease PPD incidence.

### Strengths and limitations

This project identified prepregnancy anxiety & depression as major contributors to PPD development. However, our findings do not delineate the impact of poorly controlled versus well-controlled anxiety and depression on PPD development. Prior correlational studies have suggested a linear relationship between the severity of prepregnancy anxiety and PPD risk. Non-pregnant women with higher levels of worry as defined by the Penn State Worry Questionnaire, reflecting poorly controlled anxiety, developed more PPD symptoms^18^. Our data provides perspective to these findings by demonstrating the relative importance of prepregnancy anxiety & depression in PPD development in relation to non-psychological factors like BMI, age, and income. Given the correlational relationship between control of prepregnancy psychiatric illness and PPD, our predictive models may also be used to identify expectant patients who may benefit from medications like SSRI’s; to better control prepregnancy anxiety & depression in an effort to decrease PPD incidence.

## Conclusions

PPD can be predicted with high accuracy for individual patients using antepartum information commonly found in the EMR. ML techniques offer the potential to decrease PPD incidence through early detection and subsequent CBT intervention which has been shown to decrease PPD in at-risk populations by up to 53%. Surprisingly, the presence of pre-pregnancy mental health conditions is not the most predictive factor of PPD. Our models identified BMI as the most contributing sociodemographic factor in the development of PPD. Furthermore, two of the four most contributing factors identified by our predictive models (BMI & Income) are modifiable through social policy and clinical interventions, suggesting scalable routes to decreasing the morbidity and mortality associated with PPD.

## Data Availability

All data produced are available online at

https://martinfrasch.github.io/maternal_depression_data.nb.html

## References

1. Van Niel MS, Payne JL. Perinatal depression: A review. Cleve Clin J Med. 2020;87(5):273–277. doi:10.3949/ccjm.87a.19054

2. Mughal S, Azhar Y, Siddiqui W. Postpartum Depression. In: StatPearls. Treasure Island (FL): StatPearls Publishing; 2022. https://www.ncbi.nlm.nih.gov/pubmed/30085612.

3. Jarde A, Morais M, Kingston D, et al. Neonatal Outcomes in Women With Untreated Antenatal Depression Compared With Women Without Depression: A Systematic Review and Meta-analysis. JAMA Psychiatry. 2016;73(8):826–837. doi:10.1001/jamapsychiatry.2016.0934

4. Goodman JH. Perinatal depression and infant mental health. Arch Psychiatr Nurs. 2019;33(3):217–224. doi:10.1016/j.apnu.2019.01.010

5. US Preventive Services Task Force, Curry SJ, Krist AH, et al. Interventions to Prevent Perinatal Depression: US Preventive Services Task Force Recommendation Statement. JAMA. 2019;321(6):580–587. doi:10.1001/jama.2019.0007

6. Zhang W, Liu H, Silenzio VMB, Qiu P, Gong W. Machine Learning Models for the Prediction of Postpartum Depression: Application and Comparison Based on a Cohort Study. JMIR Med Inform. 2020;8(4):e15516. doi:10.2196/15516

7. Tortajada S, García-Gomez JM, Vicente J, et al. Prediction of postpartum depression using multilayer perceptrons and pruning. Methods Inf Med. 2009;48(3):291–298. doi:10.3414/ME0562

8. Shin D, Lee KJ, Adeluwa T, Hur J. Machine Learning-Based Predictive Modeling of Postpartum Depression. J Clin Med Res. 2020;9(9). doi:10.3390/jcm9092899

9. Bennett HA, Einarson A, Taddio A, Koren G, Einarson TR. Prevalence of depression during pregnancy: systematic review. Obstet Gynecol. 2004;103(4):698–709. doi:10.1097/01.AOG.0000116689.75396.5f

10. Yang ST, Yang SQ, Duan KM, et al. The development and application of a prediction model for postpartum depression: optimizing risk assessment and prevention in the clinic. J Affect Disord. 2022;296:434–442. doi:10.1016/j.jad.2021.09.099

11. NICHD DASH - Eunice Kennedy Shriver national institute of child health and human development data and specimen hub. https://dash.nichd.nih.gov/study/226675. Accessed December 16, 2021.

12. Haas DM, Parker CB, Wing DA, et al. A description of the methods of the Nulliparous Pregnancy Outcomes Study: monitoring mothers-to-be (nuMoM2b). Am J Obstet Gynecol. 2015;212(4):539.e1-e539.e24. doi:10.1016/j.ajog.2015.01.019

13. Grobman WA, Parker CB, Willinger M, et al. Racial Disparities in Adverse Pregnancy Outcomes and Psychosocial Stress. Obstet Gynecol. 2018;131(2):328–335. doi:10.1097/AOG.0000000000002441

14. Frasch MG. Demographic and Socioeconomic Factors Predict Maternal Postpartum Rehospitalization and Depression: A Retrospective nuMoM2b Dataset Study.; 2021. doi:10.5281/zenodo.5785159

15. Surkan PJ, Sakyi KS, Christian P, et al. Risk of Depressive Symptoms Associated with Morbidity in Postpartum Women in Rural Bangladesh. Matern Child Health J. 2017;21(10):1890–1900. doi:10.1007/s10995-017-2299-7

16. Wenzel A. Basic Strategies of Cognitive Behavioral Therapy. Psychiatr Clin North Am. 2017;40(4):597–609. doi:10.1016/j.psc.2017.07.001

17. Silverman ME, Smith L, Lichtenstein P, Reichenberg A, Sandin S. The association between body mass index and postpartum depression: A population-based study. J Affect Disord. 2018;240:193–198. doi:10.1016/j.jad.2018.07.063

18. Osborne LM, Voegtline K, Standeven LR, et al. High worry in pregnancy predicts postpartum depression. J Affect Disord. 2021;294:701–706. doi:10.1016/j.jad.2021.07.009

